# Differential Prognostic Value of Angiography-Derived Microcirculatory Resistance for Coronary Artery Bypass Grafting of Intermediate Coronary Stenosis

**DOI:** 10.1101/2023.11.22.23298932

**Authors:** Wenlong Yan, Yangyang Wang, Haoyu Hu, Wei Wang, Zhenfu Li, Xin Zheng, Sumin Yang

## Abstract

**BACKGROUND:** For patients with intermediate coronary stenoses, the prognostic value of coronary artery bypass grafting (CABG) remains controversial. The prognostic impact of CABG may be different according to Murray law-based quantitative flow ratio (μQFR) and angiography-derived microcirculatory resistance (AMR) in patients with intermediate coronary stenoses.

**METHODS AND RESULTS:** Clinical data of 1411 cases who were diagnosed with multivessel coronary artery disease and had coronary artery intermediate stenoses (50% to 70% by visual assessment) between January 2013 and January 2020 were collected. Patients were categorized into CABG and defer groups based on the decision to bypass intermediate stenoses. These patients were further divided into different subgroups based on μQFR and AMR. The primary outcome was the major adverse cardiac and cerebrovascular events (MACCE), defined as a composite of cardiac death, target vessel myocardial infarction, target vessel revascularization and stroke. Multivariable analysis showed AMR (HR = 1.68, 95% CI 1.18-4.14, P = 0.008) was independent factor associated with MACCE in all patients. On receiver-operating characteristic (ROC) curve analysis, the estimated area under the curve was 0.75 (95% CI: 0.72-0.79) for MACCE. Using the maximum Youden’s index, the optimal cutoff value for AMR to predict MACCE was 2.50 mmHg*s/cm. In defer group, MACCE was significantly lower in patients with both μQFR > 0.80 and AMR < 2.50 mmHg*s/cm as compared with patients with μQFR > 0.80 (13.2% vs 22.0%, P = 0.031) or AMR < 2.50 mmHg*s/cm (13.2% vs 23.6%, P = 0.012) alone. Compared with patients with both normal μQFR and AMR, patients with both abnormal μQFR and AMR carried the highest risk for MACCE (HR = 3.46, 95% CI 2.10-5.71, P < 0.001). CABG was associated with a lower risk of MACCE compared with the deferral strategy when both μQFR and AMR were abnormal. In contrast, at normal μQFR and AMR, patients in both CABG and defer groups had similar MACCE outcomes.

**CONCLUSIONS:** In patients with intermediate coronary stenoses, the prognostic value of treatment strategy differed according to μQFR and AMR, with a significant interaction. The combination of μQFR and AMR can help stratify risk and guide treatment strategies for patients with intermediate coronary stenoses.

---

Patients with multivessel coronary artery disease experience improved survival rates after undergoing coronary artery bypass grafting (CABG), particularly those with significant comorbidities^1,2^. The standard practice for this important surgical procedure requires complete revascularization of the coronary artery. However, different studies have different definitions of complete revascularization, so the management of intermediate stenosis is still controversial^3,4^. Current guidelines recommend coronary physiologic assessment of intermediate stenoses to guide treatment in the absence of documented ischemia^5,6^. However, unlike the success of coronary physiologic assessment in percutaneous coronary intervention (PCI), it is currently believed that coronary physiologic assessment needs further research in the field of CABG^6^.

Atherosclerotic plaque-induced obstruction causes flow-limiting stenosis in an epicardial coronary artery, which is mechanistically associated with coronary artery disease (CAD). Accumulating studies demonstrate that most patients presenting with myocardial ischemia symptoms have coronary microvascular dysfunction (CMD)^7–10^. CMD affects coronary microvascular structure and/or function and was previously thought to be harmless or not harmful. However, current evidence links CMD to poor quality of life, shortened survival time, and major cardiovascular and cerebrovascular adverse events (MACCE) in CAD patients^11–13^. Equipment availability, costs, and the procedure’s long duration limit simultaneous invasive physiologic and CMD assessments. The wire- and adenosine-free angiography-derived microcirculatory resistance (AMR) index, is a valid alternative to the index of microcirculatory resistance (IMR) assessment, which is invasive and wire-based. This study assessed the clinical implications of identifying distinct disturbed coronary hemodynamic patterns using Murray law-based quantitative flow ratio (μQFR) and AMR for decision-making regarding coronary revascularization.

## METHODS

### Study population

Isolated CABG patients at the Affiliated Hospital of Qingdao University from January 2013 to January 2020 were retrospectively analyzed (Fig. 1). The inclusion criteria included: patients diagnosed with multivessel coronary artery disease and had coronary artery intermediate stenoses (50% to 70% by visual assessment). The exclusion criteria included: patients without intermediate stenoses, patients who had left main artery stenosis >50%, image data missing or with poor quality, patients who had emergency surgery, and patients with previous heart surgery. Patients were categorized into CABG and Defer groups based on the decision to bypass intermediate stenoses. The Affiliated Hospital of Qingdao University Institutional Review Board approved the study on October 30, 2023 (QYFY WZLL 28170), and waived the informed consent requirement.

**Figure1.**
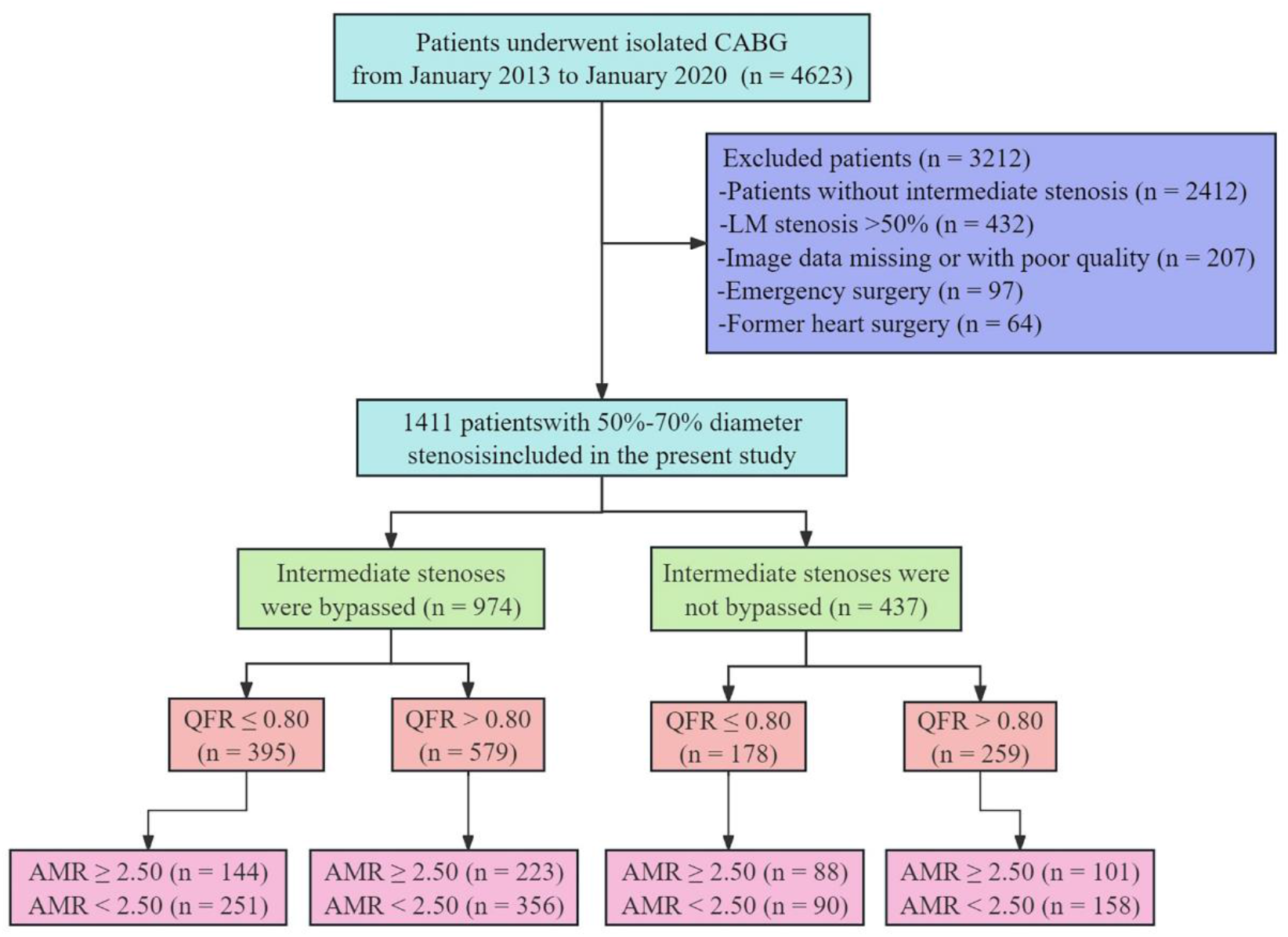
Study flow.

### Coronary angiography and physiological measurements

The coronary angiography puncture point was the radial or femoral artery, with a 5-F or 6-F sheath diameter, and 100 IU/kg intravenous heparin was used for preoperative anticoagulation. X-ray systems (Allura X per FD20, Philips; Innova IGS520, GE) were used to capture images at ≥15 frames/s. An expert certified analyst computed the single-view Murray law-based μQFR analysis using the μQFR software (AngioPlus Core, version V3, Shanghai Pulse Medical Technology Inc., Shanghai, China) at CardHemo, Med-X Research Institute, Shanghai Jiao Tong University, Shanghai, China, an independent academic core laboratory. The single-view μQFR methodology is detailed in a previous study^14^.

### Study endpoints

Telephone or outpatient follow-up was conducted, and cardiac death, target vessel myocardial infarction or revascularization, and stroke were classified as MACCE. The Academic Research Consortium report^15^ guided the clinical outcome classification and patients’ hospital records, treating cardiologist, or general practitioner confirmed adverse events.

### Statistical analysis

The analysis of normally distributed continuous data involved examining the mean standard deviation (SD) using the Student’s t-test, while for non-normally distributed continuous data, the statistical significance of the median and interquartile ranges was assessed using the Mann-Whitney U test. Categorical variables, expressed as percentages, were examined using Pearson’s χ2 or Fisher’s exact tests. Survival rates for MACCE were computed using the Kaplan-Meier analysis, and the log-rank test was used to assess the differences. Cox proportional regression analysis determined the relationship between AMR and clinical outcomes. The link between the AMR, clinical outcomes, and each variable (Table 2) was evaluated using univariate analyses. Variables with a univariate predictive value of P < 0.10 were included as covariates in the multivariable models (Table 3). The hazard ratio (HR) was estimated at a 95% confidence interval (CI). The area under the curve (AUC) of the receiver-operating characteristic (ROC) curve analysis evaluated AMR’s prediction accuracy for MACCE. Youden’s index algorithm criterion was used to identify optimal AMR cutoff values. Inverse probability of treatment weighting (IPTW) and propensity score matching (PSM) analyses were used to adjust for the uneven baseline characteristics of the CABG and Defer groups. After matching, the equilibrium was assessed using standardized mean differences (SMD). Populations were considered well-balanced if the SMD was <10%. The closest neighbor method was used to compute the PSM with a 1:2 ratio and a 0.2 caliper. R Analysis 4.2.2 (R Core Team, R Foundation for Statistical Computing, Vienna, Austria) was used for PSM and IPTW assessment. Other statistical analyses and graphics generation were performed using SPSS version 25.0 (IBM, Armonk, NY, USA). P values less than 0.05 was statistically significant.

## RESULTS

### Baseline characteristics

Figure 1 presents the study’s flowchart. Eventually, a total of 1411 patients were included, for coronary artery intermediate stenoses CABG was deferred in 437 patients (Defer group) and performed in 974 patients (CABG group). The study population’s key baseline characteristics are summarized in Table 1. About 23.2% of patients were female. The average age of the patients was 60.5 ± 8.4 years, with a BMI was 24.5 ± 3.4 kg/m2. A total of 447 (31.7%) had diabetes, 486 (34.2%) had μQFR ≤ 0.80, the mean AMR was 2.16 ± 0.82 mmHg*s/cm, and the mean SYNTAX score was 27.0 ± 10.3. The patients were followed up for a median duration of 5.9 (IQR: 3.9-8.1 years) years.

**Table 1.** Baseline characteristics of the study population.

|  | All patients (n=1411) | CABG group (n=974) | Defer group (n=437) |
| --- | --- | --- | --- |
| Age (years) | $60.5 \pm 8.4$ | $60.2 \pm 8.4$ | $61.1 \pm 8.6$ |
| Female | 327 (23.2%) | 222 (22.8%) | 105 (24.0%) |
| BMI (kg/m<sup>2</sup>) | $24.5 \pm 3.4$ | $24.5 \pm 3.5$ | $24.2 \pm 3.2$ |
| Hypertension | 866 (61.4%) | 591 (60.7%) | 275 (62.9%) |
| Diabetes mellitus | 447 (31.7%) | 302 (31.0%) | 145 (33.2%) |
| PAD | 80 (5.7%) | 50 (5.1%) | 30 (6.9%) |
| Carotid stenosis | 203 (14.4%) | 142 (14.6%) | 61 (14.0%) |
| Renal disease | 97 (6.9%) | 65 (6.7%) | 32 (7.3%) |
| Chronic lung disease | 55 (3.9%) | 39 (4.0%) | 16 (3.7%) |
| Cerebrovascular disease | 38 (2.7%) | 25 (2.6%) | 13 (3.0%) |
| Liver Disease | 46 (3.3%) | 32 (3.3%) | 14 (3.2%) |
| Smoking history | 748 (53.0%) | 509 (52.3%) | 239 (54.7%) |
| Atrial fibrillation or flutter | 26 (1.8%) | 19 (2.0%) | 7 (1.6%) |
| Prior MI | 292 (20.7%) | 206 (21.1%) | 86 (19.7%) |
| Prior PCI | 191 (13.5%) | 134 (13.8%) | 57 (13.0%) |
| LVEF<50% | 106 (7.5%) | 77 (7.9%) | 29 (6.6%) |
| NYHA class |  |  |  |
| I | 119 (8.4%) | 79 (8.1%) | 40 (9.2%) |
| II | 678 (48.1%) | 475 (48.8%) | 203 (46.5%) |
| III | 545 (38.7%) | 369 (37.9%) | 176 (40.1%) |
| IV | 69 (4.9%) | 51 (5.2%) | 18 (4.1%) |
| 3-vessel disease | 1095 (77.6%) | 767 (78.7%) | 328 (75.1%) |
| μQFR≤0.80 | 483 (34.2%) | 356 (36.6%) | 127 (29.1%) |
| AMR (mmHg*s/cm) | 2.16 ± 0.82 | 2.37 ± 0.74 | 2.01 ± 0.89 |
| SYNTAX | 27.0 ± 10.3 | 27.5 ± 10.6 | 25.9 ± 9.9 |

BMI body mass index, PAD peripheral arterial disease, MI myocardial infarction, PCI percutaneous coronary intervention, LVEF left ventricular ejection fraction, NYHA New York Heart Association, QFR quantitative flow ratio, AMR angiographic microvascular resistance

### Predictors of MACCE

Univariate analysis results showed that age, diabetes mellitus, LVEF, μQFR ≤ 0.80, AMR, and SYNTAX were relative risk factors for MACCE in all patients (Table 2). Age, diabetes mellitus, renal disease, LVEF, μQFR ≤ 0.80 and AMR were relative risk factors for MACCE in CABG group. Age, diabetes mellitus, carotid stenosis, LVEF, μQFR ≤ 0.80, AMR and SYNTAX were relative risk factors for MACCE in defer group.

**Table 2.** Univariate analysis for MACCE.

|  | All patients |  | CABG group |  | Defer group |  |
| --- | --- | --- | --- | --- | --- | --- |
|  | HR (95% CI) | P-value | HR (95% CI) | P-value | HR (95% CI) | P-value |
| Age | 1.51(1.28-2.87) | 0.005 | 1.71(1.29-3.03) | <0.001 | 1.02(1.00-1.04) | 0.035 |
| Female | 1.33(0.77-2.33) | 0.332 | 1.40(0.90-1.89) | 0.068 | 1.60(0.44-4.85) | 0.455 |
| BMI | 1.66(0.44-2.25) | 0.568 | 1.17(0.83-1.51) | 0.181 | 1.05(0.90-1.61) | 0.468 |
| Hypertension | 1.26(0.65-2.46) | 0.411 | 1.26(0.95-1.67) | 0.118 | 1.51(0.42-3.24) | 0.138 |
| Diabetes mellitus | 1.49(1.15-2.14) | 0.020 | 1.39(1.05-1.84) | 0.021 | 1.74(1.21-2.48) | 0.012 |
| PAD | 1.22(0.60-3.19) | 0.665 | 1.60(0.74-2.39) | 0.245 | 1.49(0.49-3.17) | 0.478 |
| Carotid stenosis | 1.02(0.97-1.66) | 0.268 | 1.52(0.68-3.15) | 0.411 | 1.43(1.07-2.36) | 0.021 |
| Renal disease | 1.78(0.74-4.25) | 0.194 | 1.46(1.12-3.74) | 0.025 | 1.28(0.66-3.81) | 0.098 |
| Chronic lung disease | 1.09(0.91-1.33) | 0.176 | 1.14(0.58-4.40) | 0.560 | 1.19(0.41-3.73) | 0.322 |
| Cerebrovascular disease | 1.41(0.65-3.07) | 0.375 | 1.08(0.47-3.07) | 0.131 | 1.58(0.53-4.77) | 0.478 |
| Liver Disease | 1.05(0.26-4.32) | 0.947 | 1.27(0.36-4.06) | 0.646 | 1.49(0.56-3.97) | 0.416 |
| Smoking history | 1.10(0.81-2.55) | 0.279 | 1.32(0.20-4.73) | 0.664 | 1.37(0.57-3.30) | 0.471 |
| Atrial fibrillation | 1.46(0.45-4.74) | 0.527 | 1.73(0.77-3.91) | 0.182 | 1.10(0.82-2.02) | 0.132 |
| Prior MI | 1.09(0.92-1.36) | 0.176 | 1.45(0.60-5.98) | 0.717 | 1.41(0.65-3.03) | 0.258 |
| Prior PCI | 1.10(0.89-1.66) | 0.219 | 1.53(0.77-2.18) | 0.118 | 1.24(0.74-2.08) | 0.400 |
| LVEF | 1.68(1.13-3.69) | 0.006 | 1.28(1.03-3.39) | 0.015 | 2.36(1.29-4.02) | 0.005 |
| NYHA class III IV | 1.01(0.96-1.88) | 0.132 | 1.30(0.92-1.71) | 0.090 | 1.32(0.92-1.88) | 0.124 |
| 3-vessel disease | 1.33(0.88-1.99) | 0.143 | 1.22(0.87-1.72) | 0.244 | 2.66(0.93-6.65) | 0.170 |
| μQFR≤0.80 | 1.46(1.09-4.12) | 0.005 | 1.38(1.05-2.81) | 0.008 | 1.63(1.24-2.76) | 0.018 |
| AMR | 1.79(1.22-4.68) | <0.001 | 2.11(1.18-5.87) | 0.006 | 2.54(1.28-3.41) | 0.011 |
| SYNTAX | 1.29(1.07-3.64) | 0.026 | 1.05(1.03-1.06) | 0.165 | 1.26(1.44-1.98) | <0.001 |

BMI body mass index, PAD peripheral arterial disease, MI myocardial infarction, PCI percutaneous coronary intervention, LVEF left ventricular ejection fraction, NYHA New York Heart Association, μQFR Murray law-based quantitative flow ratio, AMR angiographic microvascular resistance

In the univariate analysis, variables with P < 0.10 were used in a backward logistic regression analysis. In the multivariable analysis, age (HR = 1.21, 95% CI 1.08-1.69, P = 0.014), diabetes mellitus (HR = 1.41, 95% CI 1.10-2.37, P < 0.001), μQFR ≤ 0.80 (HR = 1.32, 95% CI 1.07-3.22, P = 0.011) and AMR (HR = 1.68, 95% CI 1.18-4.14, P = 0.008) were independent factors associated with MACCE in all patients (Table 3). Age (HR = 1.77, 95% CI 1.23-2.31, P = 0.017), LVEF (HR = 1.38, 95% CI 1.14-2.87, P = 0.002) and AMR (HR = 1.41, 95% CI 1.23-3.64, P = 0.026) were independently associated with MACCE in the CABG group. Diabetes mellitus (HR = 1.28, 95% CI 1.08-1.53, P = 0.022), μQFR ≤ 0.80 (HR = 2.11, 95% CI 1.04-4.30, P = 0.039) and AMR (HR = 1.64, 95% CI 1.11-3.69, P < 0.001) were independent predictors of MACCE in Defer group.

**Table 3.** Multivariable analysis for MACCE.

|  | All patients |  | CABG group |  | Defer group |  |
| --- | --- | --- | --- | --- | --- | --- |
|  | HR (95% CI) | P-value | HR (95% CI) | P-value | HR (95% CI) | P-value |
| Age | 1.21(1.08-1.69) | 0.014 | 1.77(1.23-2.31) | 0.017 | 1.27(0.86-1.88) | 0.225 |
| Female | - | - | 1.19(0.76-2.14) | 0.363 | - | - |
| Diabetes mellitus | 1.41(1.10-2.37) | <0.001 | 1.33(0.52-3.66) | 0.521 | 1.28(1.08-1.53) | 0.022 |
| Carotid stenosis | - | - | - | - | 1.10(0.82-1.47) | 0.525 |
| Renal disease | - | - | 1.24(0.87-1.69) | 0.274 | 1.06(0.88-1.25) | 0.470 |
| LVEF | 1.34(0.73-2.81) | 0.364 | 1.38(1.14-2.87) | 0.002 | 1.76(0.77-3.63) | 0.166 |
| NYHA class III IV | - | - | 1.38(0.83-2.11) | 0.120 | - | - |
| μQFR≤0.80 | 1.31(1.07-3.22) | 0.011 | 1.22(0.79-2.72) | 0.203 | 2.11(1.04-4.30) | 0.039 |
| AMR | 1.68(1.18-4.14) | 0.008 | 1.41(1.23-3.64) | 0.026 | 1.64(1.11-3.69) | <0.001 |
| SYNTAX | 1.19(0.94-1.99) | 0.056 | - | - | 1.53(0.74-3.74) | 0.136 |

LVEF left ventricular ejection fraction, NYHA New York Heart Association, μQFR Murray law-based quantitative flow ratio, AMR angiographic microvascular resistance

### Association between AMR and MACCE

On ROC curve analysis, the estimated AUC was 0.75 (95% CI: 0.72-0.79) for MACCE. AMR’s optimal cutoff value for predicting MACCE using Youden’s maximum index was 2.50 mmHg*s/cm (Figure 2).

**Figure 2.**
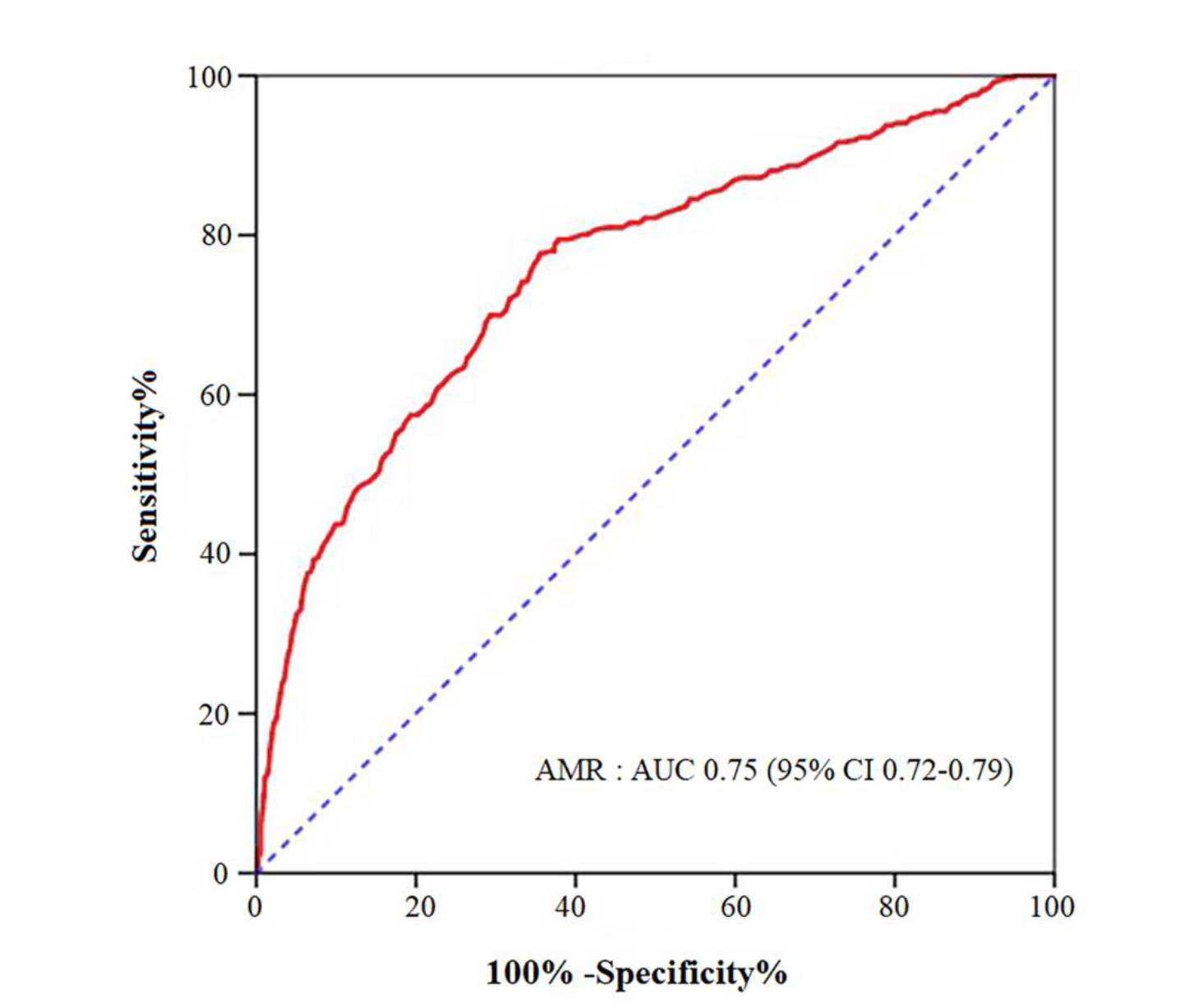
Receiver-Operating Characteristic Curve Analysis.

### Combined μQFR and AMR strategy

In defer group, we evaluated the predictive value of μQFR combined with AMR for MACCE in 437 patients. Patients with both μQFR > 0.80 and AMR < 2.50 mmHg*s/cm had significantly lower MACCE incidence than those with μQFR > 0.80 (13.2% vs 22.0%, P = 0.031) or AMR < 2.50 mmHg*s/cm (13.2% vs 23.6%, P = 0.012) alone (Figure 3).

**Figure 3.**
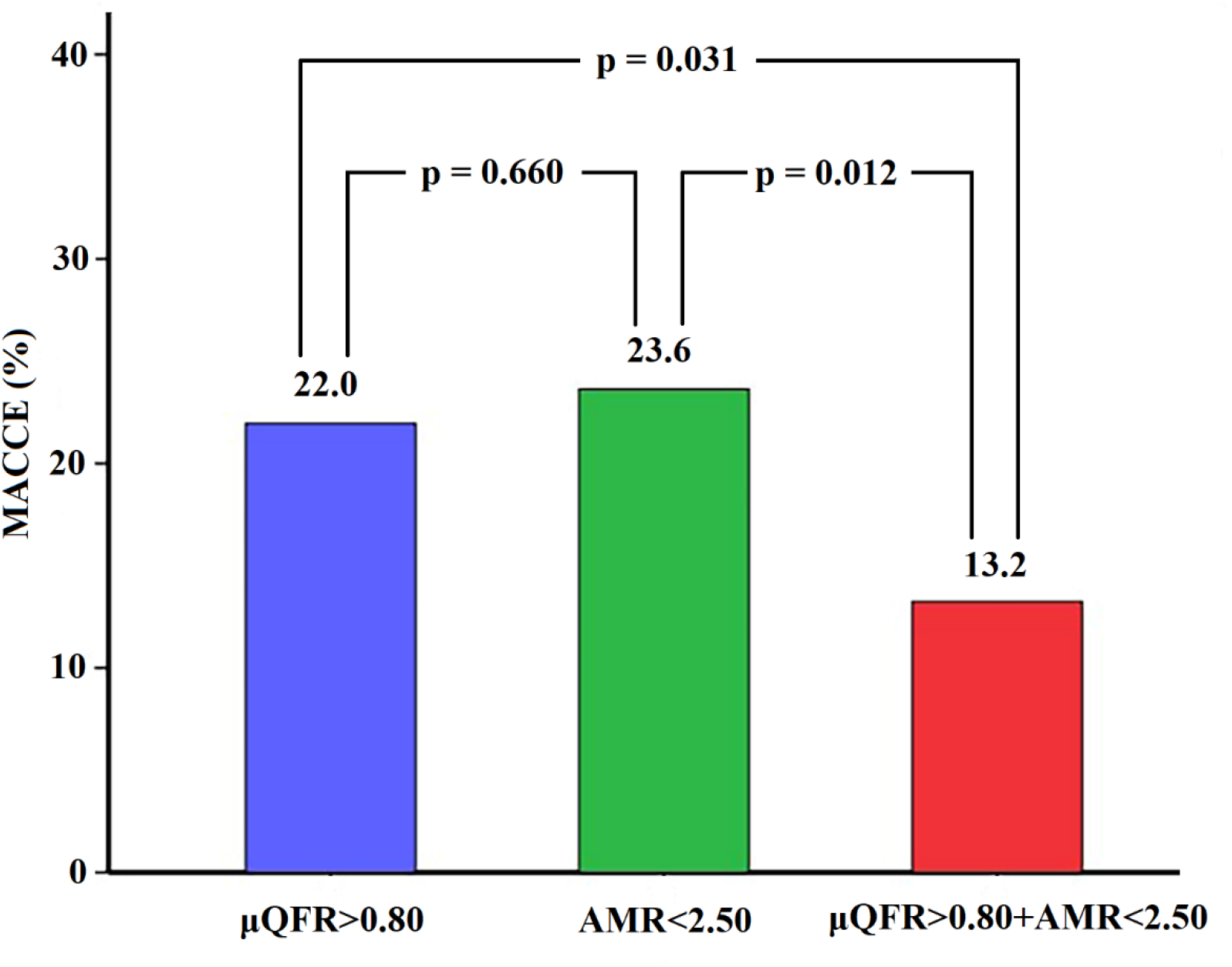
Stratified Analysis by μQFR and AMR.

### Medium-term clinical outcomes according to μQFR and AMR in defer group

The Figure 4 depicts the Kaplan-Meier curves for MACCE of 5 years across groups defined by μQFR and AMR in patients in defer group. Compared with patients with both normal μQFR and AMR, patients with both abnormal μQFR and AMR carried the highest risk for MACCE (HR = 3.46, 95% CI 2.10-5.71, P < 0.001). Discordance with normal μQFR and abnormal AMR and discordance with abnormal μQFR and normal AMR led to more MACCE than the group with normal μQFR and AMR (Group B vs Group A: HR = 2.10, 95% CI 1.23-3.59, P = 0.006; Group C vs Group A: HR = 2.43, 95% CI 1.43-4.11, P = 0.001).

**Figure 4.**
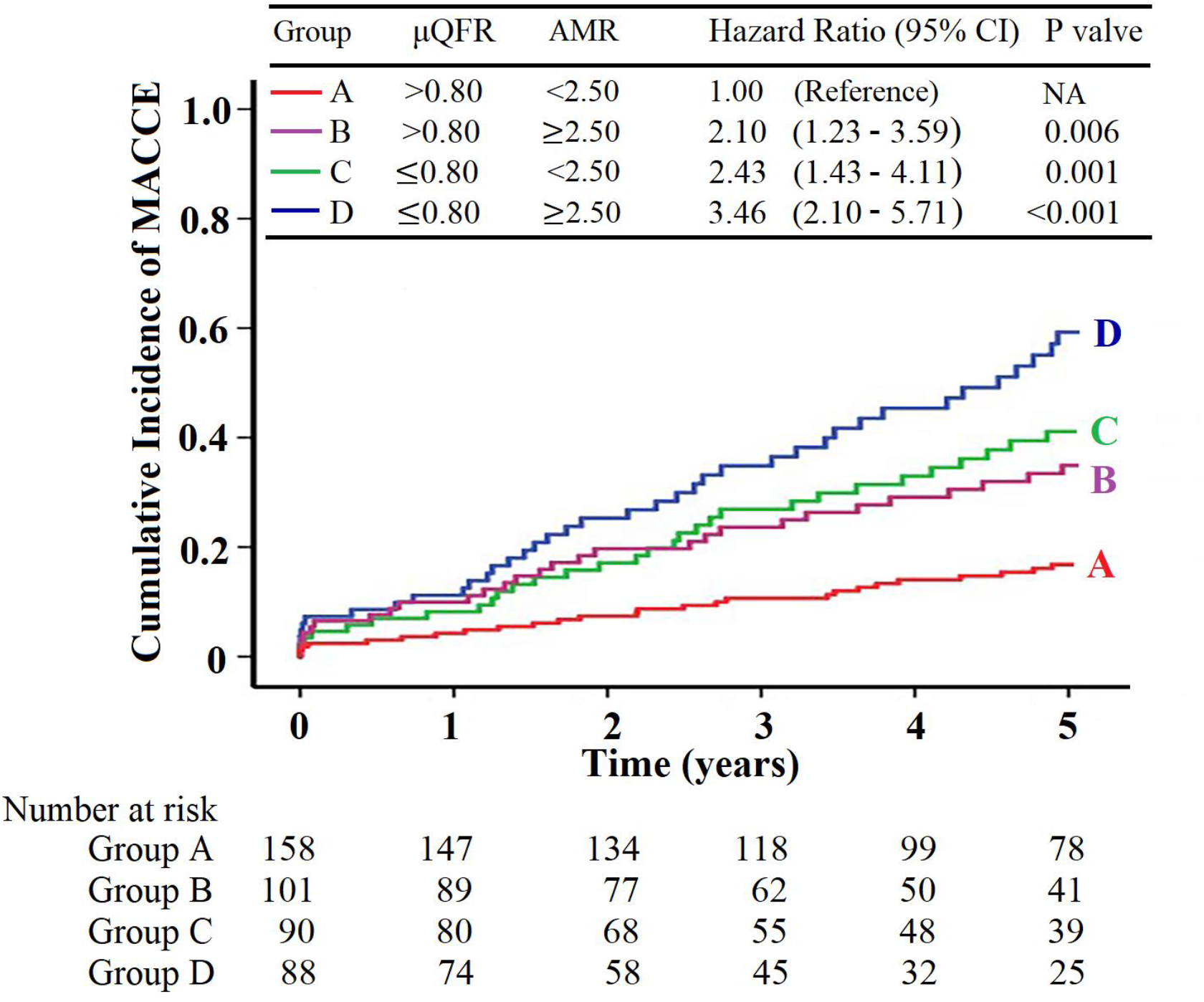
Clinical Outcomes according to μQFR and AMR in Defer Group.

### Effect of CABG on medium-term clinical outcomes divided according to μQFR and AMR

In CABG group, patients with μQFR ≤ 0.80 and AMR ≥ 2.50 mmHg*s/cm were defined as group 1 (n = 144). In defer group, patients with μQFR ≤ 0.80 and AMR ≥ 2.50 mmHg*s/cm were defined as group 2 (n = 88). The risk of MACCE was lower in group 1 than in group 2 (HR = 0.52, 95% CI 0.33-0.82, P = 0.005, Figure 5A). After PSM and IPTW matching, MACCE remained lower in group 1 than in group 2 (Figure 5B and Figure 5C).

**Figure 5.**
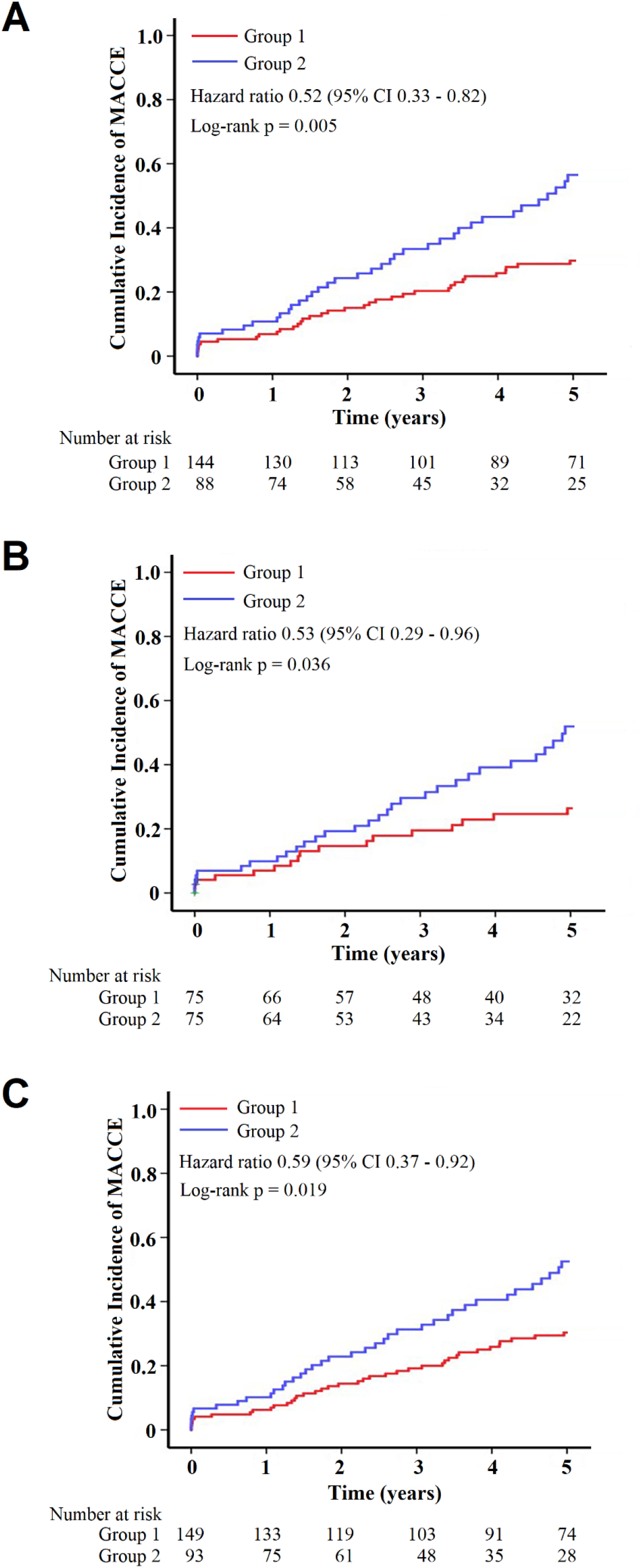
Clinical Outcomes of Group 1 and Group 2 A. Clinical Outcomes in Initial Cohort B. Clinical Outcomes of Patients after PSM C. Clinical Outcomes of Patients after IPTW.

In CABG group, patients with μQFR > 0.80 and AMR < 2.50 mmHg*s/cm were defined as group 3 (n = 356). In defer group, patients with μQFR > 0.80 and AMR < 2.50 mmHg*s/cm were defined as group 4 (n = 158). Groups 3 and 4 MACCE did not differ (HR = 1.15, 95% CI 0.73-1.80, P = 0.558, Figure 6A). After PSM and IPTW matching, groups 3 and 4 MACCE did not differ (Figure 6B and Figure 6C).

**Table 4.**
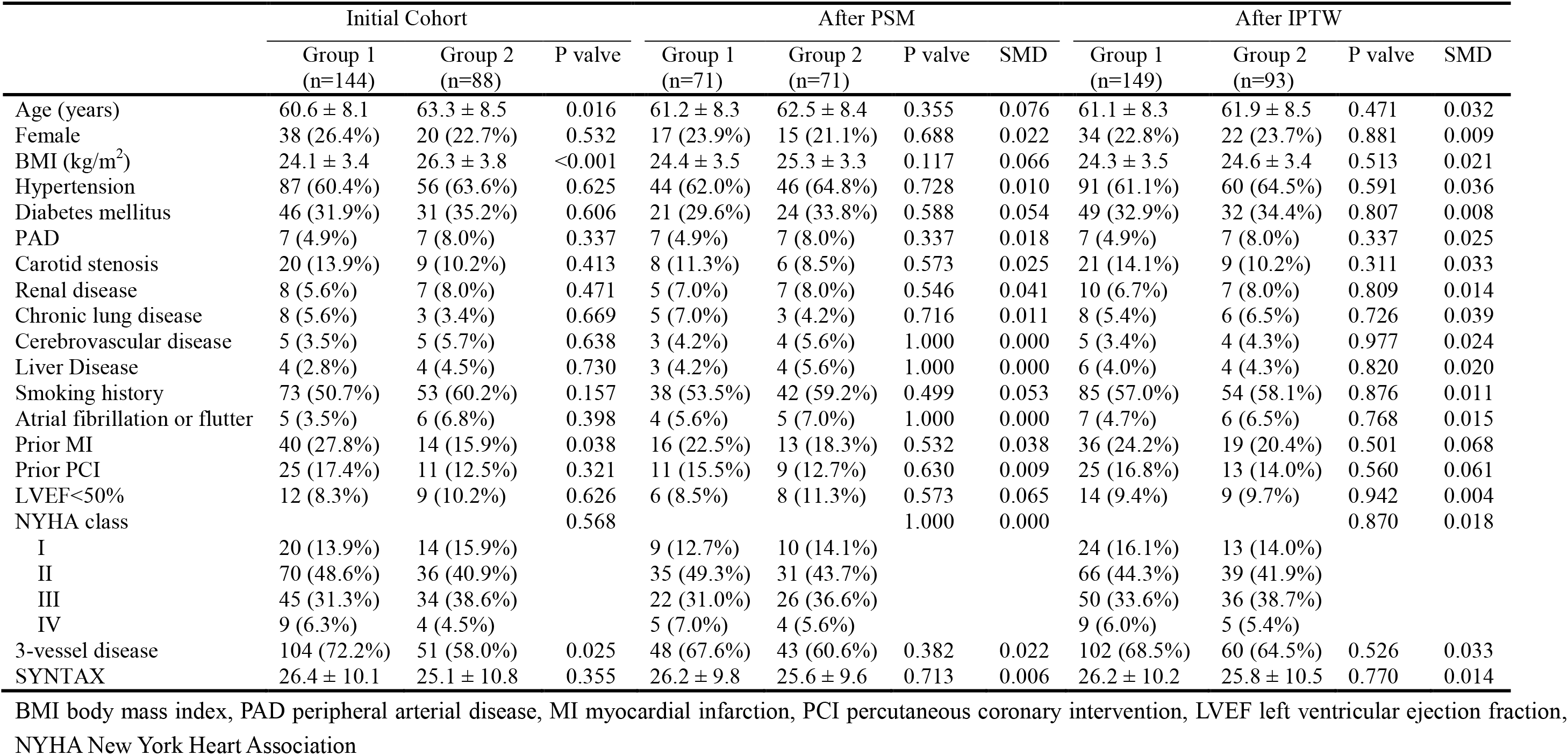
Baseline characteristics of group 1 and 2.

|  | Initial Cohort |  |  | After PSM |  |  |  | After IPTW |  |  |  |
| --- | --- | --- | --- | --- | --- | --- | --- | --- | --- | --- | --- |
|  | Group 1<br>(n=144) | Group 2<br>(n=88) | P valve | Group 1<br>(n=71) | Group 2<br>(n=71) | P valve | SMD | Group 1<br>(n=149) | Group 2<br>(n=93) | P valve | SMD |
| Age (years) | 60.6 ± 8.1 | 63.3 ± 8.5 | 0.016 | 61.2 ± 8.3 | 62.5 ± 8.4 | 0.355 | 0.076 | 61.1 ± 8.3 | 61.9 ± 8.5 | 0.471 | 0.032 |
| Female | 38 (26.4%) | 20 (22.7%) | 0.532 | 17 (23.9%) | 15 (21.1%) | 0.688 | 0.022 | 34 (22.8%) | 22 (23.7%) | 0.881 | 0.009 |
| BMI (kg/m<sup>2</sup>) | 24.1 ± 3.4 | 26.3 ± 3.8 | <0.001 | 24.4 ± 3.5 | 25.3 ± 3.3 | 0.117 | 0.066 | 24.3 ± 3.5 | 24.6 ± 3.4 | 0.513 | 0.021 |
| Hypertension | 87 (60.4%) | 56 (63.6%) | 0.625 | 44 (62.0%) | 46 (64.8%) | 0.728 | 0.010 | 91 (61.1%) | 60 (64.5%) | 0.591 | 0.036 |
| Diabetes mellitus | 46 (31.9%) | 31 (35.2%) | 0.606 | 21 (29.6%) | 24 (33.8%) | 0.588 | 0.054 | 49 (32.9%) | 32 (34.4%) | 0.807 | 0.008 |
| PAD | 7 (4.9%) | 7 (8.0%) | 0.337 | 7 (4.9%) | 7 (8.0%) | 0.337 | 0.018 | 7 (4.9%) | 7 (8.0%) | 0.337 | 0.025 |
| Carotid stenosis | 20 (13.9%) | 9 (10.2%) | 0.413 | 8 (11.3%) | 6 (8.5%) | 0.573 | 0.025 | 21 (14.1%) | 9 (10.2%) | 0.311 | 0.033 |
| Renal disease | 8 (5.6%) | 7 (8.0%) | 0.471 | 5 (7.0%) | 7 (8.0%) | 0.546 | 0.041 | 10 (6.7%) | 7 (8.0%) | 0.809 | 0.014 |
| Chronic lung disease | 8 (5.6%) | 3 (3.4%) | 0.669 | 5 (7.0%) | 3 (4.2%) | 0.716 | 0.011 | 8 (5.4%) | 6 (6.5%) | 0.726 | 0.039 |
| Cerebrovascular disease | 5 (3.5%) | 5 (5.7%) | 0.638 | 3 (4.2%) | 4 (5.6%) | 1.000 | 0.000 | 5 (3.4%) | 4 (4.3%) | 0.977 | 0.024 |
| Liver Disease | 4 (2.8%) | 4 (4.5%) | 0.730 | 3 (4.2%) | 4 (5.6%) | 1.000 | 0.000 | 6 (4.0%) | 4 (4.3%) | 0.820 | 0.020 |
| Smoking history | 73 (50.7%) | 53 (60.2%) | 0.157 | 38 (53.5%) | 42 (59.2%) | 0.499 | 0.053 | 85 (57.0%) | 54 (58.1%) | 0.876 | 0.011 |
| Atrial fibrillation or flutter | 5 (3.5%) | 6 (6.8%) | 0.398 | 4 (5.6%) | 5 (7.0%) | 1.000 | 0.000 | 7 (4.7%) | 6 (6.5%) | 0.768 | 0.015 |
| Prior MI | 40 (27.8%) | 14 (15.9%) | 0.038 | 16 (22.5%) | 13 (18.3%) | 0.532 | 0.038 | 36 (24.2%) | 19 (20.4%) | 0.501 | 0.068 |
| Prior PCI | 25 (17.4%) | 11 (12.5%) | 0.321 | 11 (15.5%) | 9 (12.7%) | 0.630 | 0.009 | 25 (16.8%) | 13 (14.0%) | 0.560 | 0.061 |
| LVEF<50% | 12 (8.3%) | 9 (10.2%) | 0.626 | 6 (8.5%) | 8 (11.3%) | 0.573 | 0.065 | 14 (9.4%) | 9 (9.7%) | 0.942 | 0.004 |
| NYHA class |  |  | 0.568 |  |  | 1.000 | 0.000 |  |  | 0.870 | 0.018 |
| I | 20 (13.9%) | 14 (15.9%) |  | 9 (12.7%) | 10 (14.1%) |  |  | 24 (16.1%) | 13 (14.0%) |  |  |
| II | 70 (48.6%) | 36 (40.9%) |  | 35 (49.3%) | 31 (43.7%) |  |  | 66 (44.3%) | 39 (41.9%) |  |  |
| III | 45 (31.3%) | 34 (38.6%) |  | 22 (31.0%) | 26 (36.6%) |  |  | 50 (33.6%) | 36 (38.7%) |  |  |
| IV | 9 (6.3%) | 4 (4.5%) |  | 5 (7.0%) | 4 (5.6%) |  |  | 9 (6.0%) | 5 (5.4%) |  |  |
| 3-vessel disease | 104 (72.2%) | 51 (58.0%) | 0.025 | 48 (67.6%) | 43 (60.6%) | 0.382 | 0.022 | 102 (68.5%) | 60 (64.5%) | 0.526 | 0.033 |
| SYNTAX | 26.4 ± 10.1 | 25.1 ± 10.8 | 0.355 | 26.2 ± 9.8 | 25.6 ± 9.6 | 0.713 | 0.006 | 26.2 ± 10.2 | 25.8 ± 10.5 | 0.770 | 0.014 |

BMI body mass index, PAD peripheral arterial disease, MI myocardial infarction, PCI percutaneous coronary intervention, LVEF left ventricular ejection fraction, NYHA New York Heart Association

**Table 5.** Baseline characteristics of group 3 and 4.

|  | Initial Cohort |  |  | After PSM |  |  |  | After IPTW |  |  |  |
| --- | --- | --- | --- | --- | --- | --- | --- | --- | --- | --- | --- |
|  | Group 3<br>(n=356) | Group 4<br>(n=158) | P valve | Group 3<br>(n=149) | Group 4<br>(n=149) | P valve | SMD | Group 3<br>(n=351) | Group 4<br>(n=162) | P<br>valve | SMD |
| Age (years) | 61.6 ± 8.8 | 60.7 ± 8.1 | 0.274 | 61.1 ± 9.3 | 60.9 ± 8.3 | 0.845 |  | 61.3 ± 9.2 | 60.8 ± 8.5 | 0.558 | 0.040 |
| Female | 72 (20.2%) | 45 (28.5%) | 0.039 | 33 (22.1%) | 39 (26.2%) | 0.417 |  | 87 (24.8%) | 43 (26.5%) | 0.671 | 0.021 |
| BMI (kg/m<sup>2</sup>) | 25.6 ± 4.2 | 24.3 ± 3.8 | 0.001 | 25.2 ± 4.1 | 24.5 ± 3.7 | 0.123 |  | 25.0 ± 4.0 | 24.6 ± 3.9 | 0.289 | 0.077 |
| Hypertension | 208 (58.4%) | 101 (63.9%) | 0.240 | 88 (59.1%) | 95 (63.8%) | 0.405 |  | 214 (61.0%) | 100 (61.7%) | 0.870 | 0.014 |
| Diabetes mellitus | 95 (26.7%) | 56 (35.4%) | 0.044 | 46 (30.9%) | 49 (32.9%) | 0.709 |  | 108 (30.8%) | 55 (34.0%) | 0.472 | 0.055 |
| PAD | 23 (6.5%) | 7 (4.4%) | 0.365 | 8 (5.4%) | 6 (4.0%) | 0.584 |  | 21 (6.0%) | 9 (5.6%) | 0.848 | 0.008 |
| Carotid stenosis | 60 (16.9%) | 18 (11.4%) | 0.111 | 21 (14.1%) | 17 (11.4%) | 0.487 |  | 52 (14.8%) | 19 (11.7%) | 0.347 | 0.024 |
| Renal disease | 17 (4.8%) | 12 (7.6%) | 0.201 | 8 (5.4%) | 10 (6.7%) | 0.627 |  | 21 (6.0%) | 11 (6.8%) | 0.725 | 0.011 |
| Chronic lung disease | 16 (4.5%) | 6 (3.8%) | 0.719 | 6 (4.0%) | 5 (3.4%) | 0.759 |  | 14 (4.0%) | 5 (3.1%) | 0.615 | 0.023 |
| Cerebrovascular disease | 11 (3.1%) | 8 (5.1%) | 0.274 | 5 (3.4%) | 7 (4.7%) | 0.556 |  | 13 (3.7%) | 7 (4.3%) | 0.737 | 0.009 |
| Liver Disease | 12 (3.4%) | 5 (3.2%) | 0.904 | 5 (3.4%) | 4 (2.7%) | 1.000 |  | 12 (3.4%) | 7 (4.3%) | 0.615 | 0.057 |
| Smoking history | 215 (60.4%) | 79 (50.0%) | 0.028 | 82 (55.0%) | 76 (51.0%) | 0.486 |  | 197 (56.1%) | 83 (51.2%) | 0.301 | 0.036 |
| Atrial fibrillation or flutter | 8 (2.2%) | 5 (3.2%) | 0.759 | 5 (3.4%) | 4 (2.7%) | 1.000 |  | 8 (2.3%) | 4 (2.5%) | 0.856 | 0.002 |
| Prior MI | 78 (21.9%) | 27 (17.1%) | 0.211 | 30 (20.1%) | 26 (17.4%) | 0.553 |  | 75 (21.4%) | 31 (19.1%) | 0.562 | 0.029 |
| Prior PCI | 45 (12.6%) | 14 (8.9%) | 0.215 | 16 (10.7%) | 13 (8.7%) | 0.558 |  | 38 (10.8%) | 16 (9.9%) | 0.745 | 0.007 |
| LVEF<50% | 28 (7.9%) | 5 (3.2%) | 0.045 | 8 (5.4%) | 4 (2.7%) | 0.239 |  | 22 (6.3%) | 8 (4.9%) | 0.551 | 0.074 |
| NYHA class |  |  | 0.518 |  |  | 0.425 |  |  |  | 0.891 | 0.001 |
| I | 39 (11.0%) | 12 (7.6%) |  | 14 (9.4%) | 10 (6.7%) |  |  | 35 (10.0%) | 13 (8.0%) |  |  |
| II | 172 (48.3%) | 77 (48.7%) |  | 83 (55.7%) | 75 (50.3%) |  |  | 169 (48.1%) | 79 (48.8%) |  |  |
| III | 124 (34.8%) | 62 (39.2%) |  | 45 (30.2%) | 58 (38.9%) |  |  | 130 (37.0%) | 63 (38.9%) |  |  |
| IV | 21 (5.9%) | 7 (4.4%) |  | 7 (4.7%) | 6 (4.0%) |  |  | 17 (4.8%) | 7 (4.3%) |  |  |
| 3-vessel disease | 223 (62.6%) | 88 (55.7%) | 0.137 | 90 (60.4%) | 84 (56.4%) | 0.481 |  | 217 (61.8%) | 95 (58.6%) | 0.493 | 0.036 |
| SYNTAX | 25.3 ± 8.8 | 23.5 ± 9.2 | 0.035 | 25.1 ± 9.1 | 24.2 ± 9.8 | 0.313 |  | 25.0 ± 9.5 | 24.6 ± 10.2 | 0.665 | 0.021 |

BMI body mass index, PAD peripheral arterial disease, MI myocardial infarction, PCI percutaneous coronary intervention, LVEF left ventricular ejection fraction, NYHA New York Heart Associa

**Figure 6.**
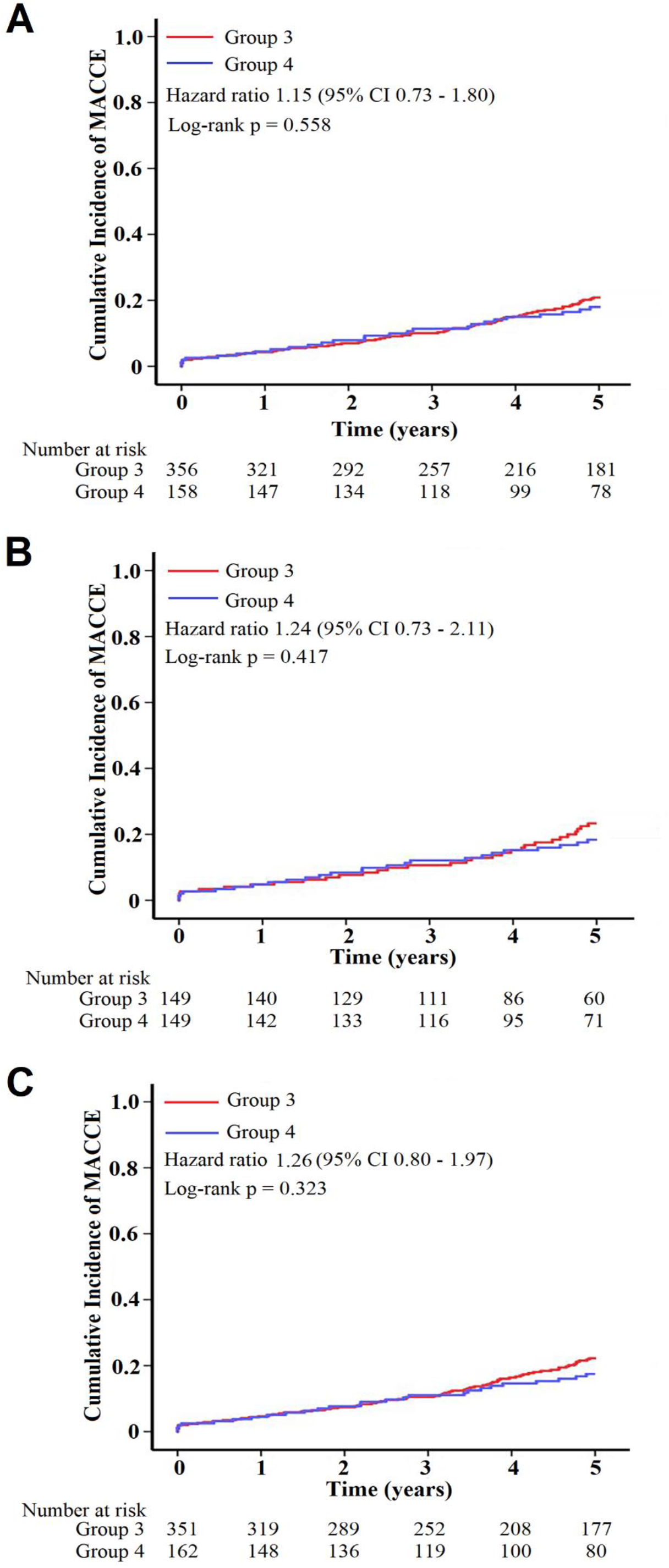
Clinical Outcomes of Group 3 and Group 4 A. Clinical Outcomes in Initial Cohort B. Clinical Outcomes of Patients after PSM C. Clinical Outcomes of Patients after IPTW.

## DISCUSSION

This study assessed the predictive significance of AMR for CABG in intermediate coronary stenosis. The primary outcomes are outlined. First, AMR independently predicted MACCE in the 5-year follow-up of patients who received CABG and those who did not. The optimal cutoff value for AMR to predict MACCE was 2.50 mmHg*s/cm, and the estimated AUC was 0.75. Second, μQFR and AMR assessments in patients with intermediate stenoses offered insights into the microvascular system that were not apparent when reviewing only the clinical or angiographic features. Patients with abnormal AMR and μQFR had worse clinical outcomes than those in other groups. Third, although CABG reduced the risk of MACCE for patients with abnormal μQFR and AMR, it did not reduce this risk for those with normal μQFR and AMR.

In ischemia with non-obstructive coronary microvascular disease (INOCA), myocardial ischemia and symptoms are caused by CMD, either alone or in combination with CAD^16^. Prior research has demonstrated that in patients without flow-limiting epicardial stenosis, microvascular disease increases the risk of cardiovascular events^17,18^. Recurrent or persistent angina, even in patients receiving effective PCI or CABG treatment, may be determined in part by CMD. In these cases, angina results from complex, multifactorial, structural, and functional causes^19^. However, hyperemic microvascular resistance (HMR) and IMR are rarely used clinically despite CMD’s prognostic potential. Their use is limited by cost, procedural time, patient discomfort from intravenous adenosine infusion, and increased procedural risk from manipulating a pressure wire in the infarcted artery. Based on computational flow analysis, several angiography-based solutions against invasive IMR have been proposed and validated for CMD investigation^20–22^. During routine angiographic evaluations, AMR demonstrated its effectiveness as a viable substitute for wire-based IMR measurements in patients with suspected or known CAD^23^. This study’s findings indicated that AMR independently predicted MACCE in all patients. The Youden test indicated that the optimal cutoff value for AMR in the ROC analysis for MACCE was 2.50 mmHg*s/cm, with an estimated AUC of 0.75 (95% CI: 0.72-0.79). This proves that AMR has good predictive value for this group of patients. We also demonstrated that lesions with a baseline AMR ≥ 2.50 mmHg*s/cm showed a higher likelihood of triggering an adverse MACCE event. AMR is easy to measure, which makes it a desirable addition to risk stratification, especially for angiographically stenotic intermediate lesions, which warrant further study.

Further, we found that μQFR combined with AMR was very good in stratifying the risk in patients with intermediate coronary stenoses who did not receive CABG. The fractional flow reserve (FFR) and μQFR can then be used to guide PCI strategies, as numerous studies have demonstrated the relationship between coronary physiologic assessment indices and clinical outcomes^24–26^. Based on these insights, it has been suggested that FFR and μQFR could be used to guide target arteries for CABG. Finally, some researchers believe that CABG prolongs life expectancy, and using FFR to direct revascularization is not as logical for CABG as it is for PCI^27^. Our study explored the guiding value of coronary artery physiologic assessment indices in CABG from a new perspective. For patients with intermediate coronary stenoses who did not receive CABG, those with both normal μQFR and AMR had significantly lower MACCE than those with normal μQFR or AMR alone (Figure 3). Meanwhile, compared with both normal μQFR and AMR (Group A), both abnormal μQFR and AMR (Group D) carried the highest risk for MACCE (Figure 4). After finding this interesting result, we compared the effects of CABG on these different risk groups. For patients with both abnormal μQFR and AMR, MACCE reduced significantly in the CABG group compared to the Defer group. Conversely, for patients with both normal μQFR and AMR, both groups had a similar risk of MACCE. And the results were consistent in the initial patients and patients after PSM or IPTW matching (Figures 5 6). These results demonstrate that in patients with intermediate coronary stenoses, CABG is beneficial in the cohort with both abnormal μQFR and AMR cohort but not in those with normal μQFR and AMR. This may be explained by the improved microvascular resistance in the abnormal μQFR and AMR cohorts due to CABG-induced epicardial coronary stenosis relief, leading to increased myocardial perfusion. Despite the widespread belief that PCI or CABG do not affect microvascular resistance, a previous study demonstrated that after coronary revascularization, increased distal coronary pressure significantly reduced the microvascular resistance index^28^. Similarly, findings from the current study indicate that combining μQFR with AMR can effectively identify patients with intermediate coronary stenoses who may benefit from CABG and those who could be safely deferred.

### Study limitations

First, this study has the inherent drawbacks of a retrospective registry, even with the large sample size and follow-up duration. As a result, a prospective randomized study is required to confirm these conclusions. Second, AMR relies on angiography, image quality, and optimal projection. For this study’s AMR analysis, we excluded 4.5% of the patients because the angiographic image was of low quality. Third, selection bias may have occurred because the final decision for CABG was at the operator’s discretion. Although the results of the multivariable adjusted analysis were consistent with the PSM and IPTW analysis, it is important to take into account the potential impact of unmeasured bias.

## CONCLUSIONS

In the 5-year-follow-up evaluation, AMR independently predicted MACCE in patients with intermediate coronary stenoses. The treatment strategy’s prognostic values varied, with significant interaction, based on the μQFR and AMR. CABG was associated with a lower risk of MACCE compared with the deferral strategy when both μQFR and AMR were abnormal. In contrast, at normal μQFR and AMR, patients in both CABG and Defer groups had similar MACCE outcomes. The combination of μQFR and AMR can help stratify risk and guide treatment strategies for patients with intermediate coronary stenoses.

## Data Availability

The authors will supply the relevant data in response to reasonable requests.

## References

1. Bianco V, Kilic A, Mulukutla S, Gleason TG, Kliner D, Allen CC, Habertheuer A, Aranda-Michel E, Humar R, Navid F, et al. Percutaneous coronary intervention versus coronary artery bypass grafting in patients with reduced ejection fraction. J Thorac Cardiovasc Surg. 2021;161. doi: 10.1016/j.jtcvs.2020.06.159

2. Holm NR, Mäkikallio T, Lindsay MM, Spence MS, Erglis A, Menown IBA, Trovik T, Kellerth T, Kalinauskas G, Mogensen LJH, et al. Percutaneous coronary angioplasty versus coronary artery bypass grafting in the treatment of unprotected left main stenosis: updated 5-year outcomes from the randomised, non-inferiority NOBLE trial. Lancet. 2020;395:191–199. doi: 10.1016/S0140-6736(19)32972-1

3. Gaba P, Gersh BJ, Ali ZA, Moses JW, Stone GW. Complete versus incomplete coronary revascularization: definitions, assessment and outcomes. Nat Rev Cardiol. 2021;18:155–168. doi: 10.1038/s41569-020-00457-5

4. Leviner DB, Torregrossa G, Puskas JD. Incomplete revascularization: what the surgeon needs to know. Ann Cardiothorac Surg. 2018;7:463–469. doi: 10.21037/acs.2018.06.07

5. Neumann F-J, Sousa-Uva M, Ahlsson A, Alfonso F, Banning AP, Benedetto U, Byrne RA, Collet J-P, Falk V, Head SJ, et al. 2018 ESC/EACTS Guidelines on myocardial revascularization. Eur Heart J. 2019;40. doi: 10.1093/eurheartj/ehy394

6. Lawton JS, Tamis-Holland JE, Bangalore S, Bates ER, Beckie TM, Bischoff JM, Bittl JA, Cohen MG, DiMaio JM, Don CW, et al. 2021 ACC/AHA/SCAI Guideline for Coronary Artery Revascularization: A Report of the American College of Cardiology/American Heart Association Joint Committee on Clinical Practice Guidelines. J Am Coll Cardiol. 2022;79. doi: 10.1016/j.jacc.2021.09.006

7. Ya’qoub L, Elgendy IY, Pepine CJ. Syndrome of Nonobstructive Coronary Artery Diseases: A Comprehensive Overview of Open Artery Ischemia. Am J Med. 2021;134:1321–1329. doi: 10.1016/j.amjmed.2021.06.038

8. Beltrame JF, Tavella R, Jones D, Zeitz C. Management of ischaemia with non-obstructive coronary arteries (INOCA). BMJ. 2021;375:e060602. doi: 10.1136/bmj-2021-060602

9. Kunadian V, Chieffo A, Camici PG, Berry C, Escaned J, Maas AHEM, Prescott E, Karam N, Appelman Y, Fraccaro C, et al. An EAPCI Expert Consensus Document on Ischaemia with Non-Obstructive Coronary Arteries in Collaboration with European Society of Cardiology Working Group on Coronary Pathophysiology & Microcirculation Endorsed by Coronary Vasomotor Disorders International Study Group. Eur Heart J. 2020;41:3504–3520. doi: 10.1093/eurheartj/ehaa503

10. Maddox TM, Stanislawski MA, Grunwald GK, Bradley SM, Ho PM, Tsai TT, Patel MR, Sandhu A, Valle J, Magid DJ, et al. Nonobstructive coronary artery disease and risk of myocardial infarction. JAMA. 2014;312:1754–1763. doi: 10.1001/jama.2014.14681

11. Taqueti VR, Solomon SD, Shah AM, Desai AS, Groarke JD, Osborne MT, Hainer J, Bibbo CF, Dorbala S, Blankstein R, et al. Coronary microvascular dysfunction and future risk of heart failure with preserved ejection fraction. Eur Heart J. 2018;39:840–849. doi: 10.1093/eurheartj/ehx721

12. Schumann CL, Mathew RC, Dean J-HL, Yang Y, Balfour PC, Shaw PW, Robinson AA, Salerno M, Kramer CM, Bourque JM. Functional and Economic Impact of INOCA and Influence of Coronary Microvascular Dysfunction. JACC Cardiovasc Imaging. 2021;14:1369–1379. doi: 10.1016/j.jcmg.2021.01.041

13. Del Buono MG, Montone RA, Camilli M, Carbone S, Narula J, Lavie CJ, Niccoli G, Crea F. Coronary Microvascular Dysfunction Across the Spectrum of Cardiovascular Diseases: JACC State-of-the-Art Review. J Am Coll Cardiol. 2021;78:1352–1371. doi: 10.1016/j.jacc.2021.07.042

14. Tu S, Ding D, Chang Y, Li C, Wijns W, Xu B. Diagnostic accuracy of quantitative flow ratio for assessment of coronary stenosis significance from a single angiographic view: A novel method based on bifurcation fractal law. Catheter Cardiovasc Interv. 2021;97 Suppl 2:1040–1047. doi: 10.1002/ccd.29592

15. Garcia-Garcia HM, McFadden EP, Farb A, Mehran R, Stone GW, Spertus J, Onuma Y, Morel M-A, van Es G-A, Zuckerman B, et al. Standardized End Point Definitions for Coronary Intervention Trials: The Academic Research Consortium-2 Consensus Document. Eur Heart J. 2018;39:2192–2207. doi: 10.1093/eurheartj/ehy223

16. Ong P, Camici PG, Beltrame JF, Crea F, Shimokawa H, Sechtem U, Kaski JC, Bairey Merz CN. International standardization of diagnostic criteria for microvascular angina. Int J Cardiol. 2018;250:16–20. doi: 10.1016/j.ijcard.2017.08.068

17. Dhawan SS, Corban MT, Nanjundappa RA, Eshtehardi P, McDaniel MC, Kwarteng CA, Samady H. Coronary microvascular dysfunction is associated with higher frequency of thin-cap fibroatheroma. Atherosclerosis. 2012;223:384–388. doi: 10.1016/j.atherosclerosis.2012.05.034

18. Fearon WF, Low AF, Yong AS, McGeoch R, Berry C, Shah MG, Ho MY, Kim H-S, Loh JP, Oldroyd KG. Prognostic value of the Index of Microcirculatory Resistance measured after primary percutaneous coronary intervention. Circulation. 2013;127:2436–2441. doi: 10.1161/CIRCULATIONAHA.112.000298

19. Mangiacapra F, Del Buono MG, Abbate A, Gori T, Barbato E, Montone RA, Crea F, Niccoli G. Role of endothelial dysfunction in determining angina after percutaneous coronary intervention: Learning from pathophysiology to optimize treatment. Prog Cardiovasc Dis. 2020;63:233–242. doi: 10.1016/j.pcad.2020.02.009

20. Scarsini R, Shanmuganathan M, Kotronias RA, Terentes-Printzios D, Borlotti A, Langrish JP, Lucking AJ, Ribichini F, Ferreira VM, Channon KM, et al. Angiography-derived index of microcirculatory resistance (IMRangio) as a novel pressure-wire-free tool to assess coronary microvascular dysfunction in acute coronary syndromes and stable coronary artery disease. Int J Cardiovasc Imaging. 2021;37:1801–1813. doi: 10.1007/s10554-021-02254-8

21. De Maria GL, Scarsini R, Shanmuganathan M, Kotronias RA, Terentes-Printzios D, Borlotti A, Langrish JP, Lucking AJ, Choudhury RP, Kharbanda R, et al. Angiography-derived index of microcirculatory resistance as a novel, pressure-wire-free tool to assess coronary microcirculation in ST elevation myocardial infarction. Int J Cardiovasc Imaging. 2020;36:1395–1406. doi: 10.1007/s10554-020-01831-7

22. Tebaldi M, Biscaglia S, Di Girolamo D, Erriquez A, Penzo C, Tumscitz C, Campo G. Angio-Based Index of Microcirculatory Resistance for the Assessment of the Coronary Resistance: A Proof of Concept Study. J Interv Cardiol. 2020;2020:8887369. doi: 10.1155/2020/8887369

23. Fan Y, Fezzi S, Sun P, Ding N, Li X, Hu X, Wang S, Wijns W, Lu Z, Tu S. In Vivo Validation of a Novel Computational Approach to Assess Microcirculatory Resistance Based on a Single Angiographic View. J Pers Med. 2022;12. doi: 10.3390/jpm12111798

24. Zimmermann FM, Omerovic E, Fournier S, Kelbæk H, Johnson NP, Rothenbühler M, Xaplanteris P, Abdel-Wahab M, Barbato E, Høfsten DE, et al. Fractional flow reserve-guided percutaneous coronary intervention vs. medical therapy for patients with stable coronary lesions: meta-analysis of individual patient data. Eur Heart J. 2019;40:180–186. doi: 10.1093/eurheartj/ehy812

25. Xaplanteris P, Fournier S, Pijls NHJ, Fearon WF, Barbato E, Tonino PAL, Engstrøm T, Kääb S, Dambrink J-H, Rioufol G, et al. Five-Year Outcomes with PCI Guided by Fractional Flow Reserve. N Engl J Med. 2018;379:250–259. doi: 10.1056/NEJMoa1803538

26. Xu B, Tu S, Song L, Jin Z, Yu B, Fu G, Zhou Y, Wang Ja, Chen Y, Pu J, et al. Angiographic quantitative flow ratio-guided coronary intervention (FAVOR III China): a multicentre, randomised, sham-controlled trial. Lancet. 2021;398:2149–2159. doi: 10.1016/S0140-6736(21)02248-0

27. Piccolo R, Giustino G, Mehran R, Windecker S. Stable coronary artery disease: revascularisation and invasive strategies. Lancet. 2015;386:702–713. doi: 10.1016/S0140-6736(15)61220-X

28. Verhoeff B-J, Siebes M, Meuwissen M, Atasever B, Voskuil M, de Winter RJ, Koch KT, Tijssen JGP, Spaan JAE, Piek JJ. Influence of percutaneous coronary intervention on coronary microvascular resistance index. Circulation. 2005;111:76–82.

